# Weight loss with a lower dose compounded semaglutide and behavioral weight management program: A real-world matched retrospective cohort study

**DOI:** 10.64898/2026.09.08.26362528

**Authors:** Erin C. Owen, Robert G. Moulder, Jessica L. Morse, M. Cole Ainsworth, Alexander Fabry, Amanda R. Merner

## Abstract

**Objective:** This 16-week, real-world retrospective cohort study of a behavioral weight management program evaluated the efficacy of a lower dose of compounded semaglutide on weight loss.

**Methods:** Weight loss was compared among Noom Weight (NW) users, Noom Microdose GLP-1 Rx Program (NMGP) users prescribed a low dose of compounded semaglutide (up to 0.6mg/weekly), and Noom GLP-1 Rx Program (NGP) users prescribed a dose up to 1.2mg/weekly of compounded semaglutide via multiple-group latent growth curve models for the full sample and propensity score matched cohort. Users were matched on demographic characteristics, Area Deprivation Index, baseline Body Mass Index, and urbanicity.

**Results:** In the full sample, participants (N = 29,788) were mostly middle-aged, urban-dwelling females. At 16 weeks, NW users lost 2.6%, NMGP users lost 8.3%, and NGP users lost 8.6% of their baseline weight (all comparisons *p* ≤ .001). In the matched cohort (N = 3,657; n= 1,219 per group), participant characteristics were similar. At 16 weeks, NW users lost 2.6%, NMGP users lost 8.3%, and NGP users lost 9.1% of their initial weight (all comparisons *p* < .001).

**Conclusions:** This large, retrospective cohort reports lower doses of compounded semaglutide alongside a behavioral companion achieve 91% of the weight loss of a higher dose of compounded semaglutide in the first 16 weeks.

## Introduction

Obesity is a prevalent and significant public health concern. Estimates suggest 40.3% of U.S. adults met criteria for obesity in 2025 and nearly 75% met criteria for overweight or obesity (1). Obesity is associated with an increased risk of hypertension and hypercholesterolemia, obstructive sleep apnea, various forms of cancer and cardiovascular disease, and is linked to approximately 50% of new Type 2 Diabetes cases in the U.S. annually (1). Risk of mortality is heightened among individuals with obesity, with a 30% higher risk of death with every five-point increase in body mass index (BMI) above 25 (1). In addition to its impacts on health, obesity imposes a significant economic burden, costing the U.S. approximately $175 billion annually (1). An estimated 12% of adults in the United States use GLP-1 medications (2), of which 19% report using compounded formulations (3). Despite the relatively high percentage of compounded users, almost all published trials of GLP-1s have examined FDA approved formulations. Studies examining real-world use of compounded semaglutide medications present an opportunity to advance understanding of safety, tolerability, and efficacy.

It is well established that FDA approved semaglutide supports meaningful weight loss (4). Although GLP-1s appear promising in terms of weight loss, the financial costs and side effect profiles associated with these medications can impede treatment (2). For those without insurance coverage, higher dosages typically cost more (5). In addition to financial costs, side effects can negatively impact health and wellbeing. Real-world studies reveal discontinuation rates at 12 months ranging from 45.2% to 64.8% (4,6). By 36 months, research reveals over 90% of users discontinue GLP-1 medications (7).

One approach to reducing cost and side effect barriers associated with initiating and continuing use of GLP-1s is prescribing lower doses of these medications. Noom is a digital health company that offers app-based behavioral weight management programs alongside clinician-supervised treatment with compounded semaglutide, when medically appropriate. Noom’s definition of a lower dose is 25% or less of the standard commercially available dosage of FDA approved semaglutide, 2.4g weekly (8). Lower doses may be less expensive, which reduces cost-related barriers to treatment, and studies reveal clinically meaningful weight loss on doses below FDA-approved maximums (9–11). Research has shown an average weight loss over three months with lower doses, including weight loss of 4.8kg on 1.6mg/weekly FDA approved semaglutide (11). Weight loss of 5% or greater is considered clinically meaningful, thus doses significantly lower than FDA-approved doses, may promote meaningful weight loss and be more tolerable. Dose-ranging studies also reveal a clear dose-response on side effects (11). A dose-dependent increase in gastrointestinal (GI) adverse events (e.g., nausea, vomiting) has been noted with semaglutide (11), and GI side effects observed with higher doses of semaglutide have been deemed unacceptable for continued administration by some (11). Overall, evidence supports dose-dependent side effects and weight loss for branded semaglutide.

The vast majority of GLP-1 RA users will not be lifetime users. However, significant weight regain has been observed upon discontinuation of standard dose of FDA approved GLP-1 medications, with an average monthly rate just below 1lb and a return of cardiometabolic markers to baseline within 1.5 years of GLP-1 RA cessation (see (12) for review). Weight regain appears to occur almost four times faster after GLP-1 RA discontinuation compared to behavioral weight management program cessation (13), which may perpetuate reinitiation of GLP-1s (4) and weight cycling. Behavioral weight management programs incorporate education, support, and skills for individuals seeking to increase physical activity, improve diet, and address psychological factors that contribute to unhealthy behaviors (14).

Pairing a GLP-1 RA with a behavioral weight management program may improve initial weight loss, long-term weight maintenance, and overall health. A 68-week RCT of FDA approved semaglutide compared to placebo in conjunction with intensive behavioral therapy (STEP 3) resulted in reductions in body weight of 16% versus 5.7%, respectively (15). A real-world study of a digital health program combined with personalized doses of FDA approved semaglutide (Embla ApS) revealed average weight loss of 16.7% after 64 weeks with a mean semaglutide dose of 1.08mg/weekly (9), further substantiating (11) that doses lower than the standard, or FDA-approved maximum dose, may be effective in producing clinically meaningful weight loss and that combining the two (medication and behavioral tools) may be particularly effective for long-term weight loss.

While many studies have separately examined weight loss for participants enrolled in behavioral weight management programs and users of standard doses of FDA approved semaglutide, weight loss effects of lower doses of semaglutide while enrolled in behavioral weight management have been minimally studied (9). This approach may hold promise for clinically significant weight loss with potentially fewer side effects. In the present study, we examined retrospective data collected over the first 16 weeks of enrollment in Noom’s weight management programs, which are grounded in cognitive behavioral principles that incorporate psychoeducation, self-monitoring, human coaching, and community support. We examined weight loss among Noom users taking a lower dose of compounded semaglutide in conjunction with a behavioral program (Noom Microdose GLP-1 Rx Program; NMGP) to determine if it produces clinically significant, short-term weight loss (i.e., 16 weeks) and compared NMGP participants to participants taking Noom’s standard dose of a compounded semaglutide (Noom GLP-1 Rx Program; NGP) and unmedicated Noom Weight users (NW) (Aim 1). We then matched participants across programs on demographic, socioeconomic, and clinical characteristics and conducted the same weight comparisons (Aim 2).

## Methods

### Study Design

This matched-cohort retrospective study used a dataset of participants new to Noom who enrolled in either NW, NMGP, or NGP programs between July 21, 2025, and August 31, 2025 and were followed through December 31, 2025. This date range reflects Noom users who enrolled in NMGP at launch and were followed for the first 16 weeks. All procedures were approved by the Advarra institutional review board (Protocol Number: Pro00076674). For matched analyses, NW, NMGP, or NGP participants were matched on the following characteristics: sex, age, baseline BMI, region of the United States, and living environment (rural or urban). In addition, participants were matched on Area Deprivation Index (ADI), a measure of how socioeconomically deprived residents of a geographic area are, on average, and is considered a measure of social risk strongly associated with health and mortality (16).

### Participants

All participants were required to be US residents, aged 18 years to 80 years and enrolled in one of three Noom programs (NW, NMGP, or NGP) with a paid subscription after program-specific trial periods (two weeks for NW and three weeks for NMGP and NGP). Participants were excluded from NW if they had a history of an eating disorder. Participants were excluded from NMGP and NGP if they had: BMI < 25, active cancer, liver failure, severe heart disease, personal or family history of Multiple Endocrine Neoplasia Type 2, personal or family history of Medullary Thyroid Cancer, benzoyl alcohol allergy, were pregnant or nursing, or reside in a state that NMGP or NGP do not operate. Additional exclusion criteria for this study included a diagnosis of Type 2 diabetes and program participants who switched programs during the four-month study period.

Study attrition was defined as the percentage of participants who did not report their weight at each time point, representing a proxy for potential program attrition. Direct measures of program discontinuation (e.g., dropout, not refilling medications) were not captured in the scope of the present study. At baseline, 70.2% of participants reported their weight, at week 4, 70.3% reported their weight, at week 8, 56.0% reported their weight, at week 12, 44.5% reported their weight, and at week 16, 32.5% reported their weight.

### Measures

#### Demographics

Demographic variables, including gender, age, and BMI, were self-reported at program onboarding. Geographic region and urbanicity were derived from the participant’s residential zip code using Rural-Urban Commuting Area (RUCA) codes. Zip Code Tabulation Area (ZCTA) was used to derive Area Deprivation Index (ADI).

#### Weight

Participants self-reported their body weight (lbs) as part of routine engagement in the Noom application, which prompts for daily weight logging. For participants in NMGP and NGP, weight was also captured in the electronic health record in advance of clinical appointments. This study utilized self-reported weight logged due to the number of observations available.

### Programs

#### NW

NW is a commercially available, mobile app focused on lifestyle, nutrition, and weight loss via behavior change techniques based on principles of cognitive behavioral therapy (CBT), acceptance and commitment therapy (ACT), and dialectical behavior therapy (DBT). NW offers users various engagement options, with a focus on self-monitoring (e.g., nutrition, weight, physical activity) and education (e.g., articles) to change patterns. Previous work describes NW in greater detail (17) as well as NW’s effectiveness as a behavioral weight intervention (18).

#### GLP-1 Rx Programs: NGP and NMGP

Individuals who signed up for Noom and expressed an interest in prescription weight loss medications were required to complete a medical intake and clinical review to determine eligibility with a licensed medical provider contracted through Noom. Across both GLP-1 Rx programs (NMGP and NGP), users titrated doses of compounded semaglutide slowly to mitigate against side effects and avoid too rapid of weight loss. Clinical factors such as history of GLP-1 use and associated experience influenced baseline dosing. These Noom programs utilize personalized dosing along with medical care (i.e., routine check-ins and 24/7 medical provider access) to accommodate users’ needs, weight loss trajectories, side effects, and more, adjusting dosages as needed. In addition to medication and medication management (e.g., medication logging, side effect logging, chat options with medical providers), NMGP and NGP users received a Success Kit, which provided guidance on avoidance and management of side effects while titrating GLP-1 doses over time. Users were also enrolled in the Noom companion experience, which includes the core features in the NW app as well as an exercise library curated for muscle preservation and a psychology-based curriculum that encourages mindful eating practices, provides meal guidance and AI-based food logging with protein tracking.

#### Compounded Semaglutide Manufacturing and Dosing Protocols

Noom’s compounding pharmacy partners operate under Section 503A of the Federal Food, Drug, and Cosmetic Act, 21 U.S.C. § 353a (19,20). With oversight sitting primarily with the State Boards of Pharmacy, Noom’s compounding pharmacies must adhere to United States Pharmacopeia (USP) guidelines, and every batch of medication is tested to ensure quality and safety. The compounded semaglutide formulation consists of semaglutide (2.5 mg/mL) and glycine, USP (5 mg/mL), in a vehicle containing sodium phosphate buffer, propylene glycol, USP (1.5% v/v), and phenol (0.55%) as a preservative, with pH adjusted using sodium hydroxide and/or hydrochloric acid as needed. Water for Injection, USP, was added quantum sufficiat (q.s.) to the final volume. Noom NMGP and NGP participants may request a Certificate of Analysis provided by the compounding pharmacy.

The dosing protocol of compounded semaglutide for NMGP and NGP are identical for the first eight weeks (starting at 0.2mg/week and titrating up to 0.6mg/week), although allowed to vary as needed based on side effect tolerance and weight loss progress. After week 8, the dosing protocol for NMGP allows for a maximum dose of up to 0.6mg/weekly compounded semaglutide. The NGP protocol allows for an increase in dose to 0.85mg/weekly during weeks nine and ten and to up to 1.2mg/weekly during weeks 11 and beyond. Thus, the maximum dose for NMGP participants is 0.6mg/weekly and the maximum dose for NGP participants is 1.2mg/weekly. To confirm whether the protocols accurately reflect real-world titration experiences under Noom’s personalized dosing approach, we calculated a violin plot showing the distribution of actual doses taken by participants in each program per week (Figure 1).

**Figure 1.**
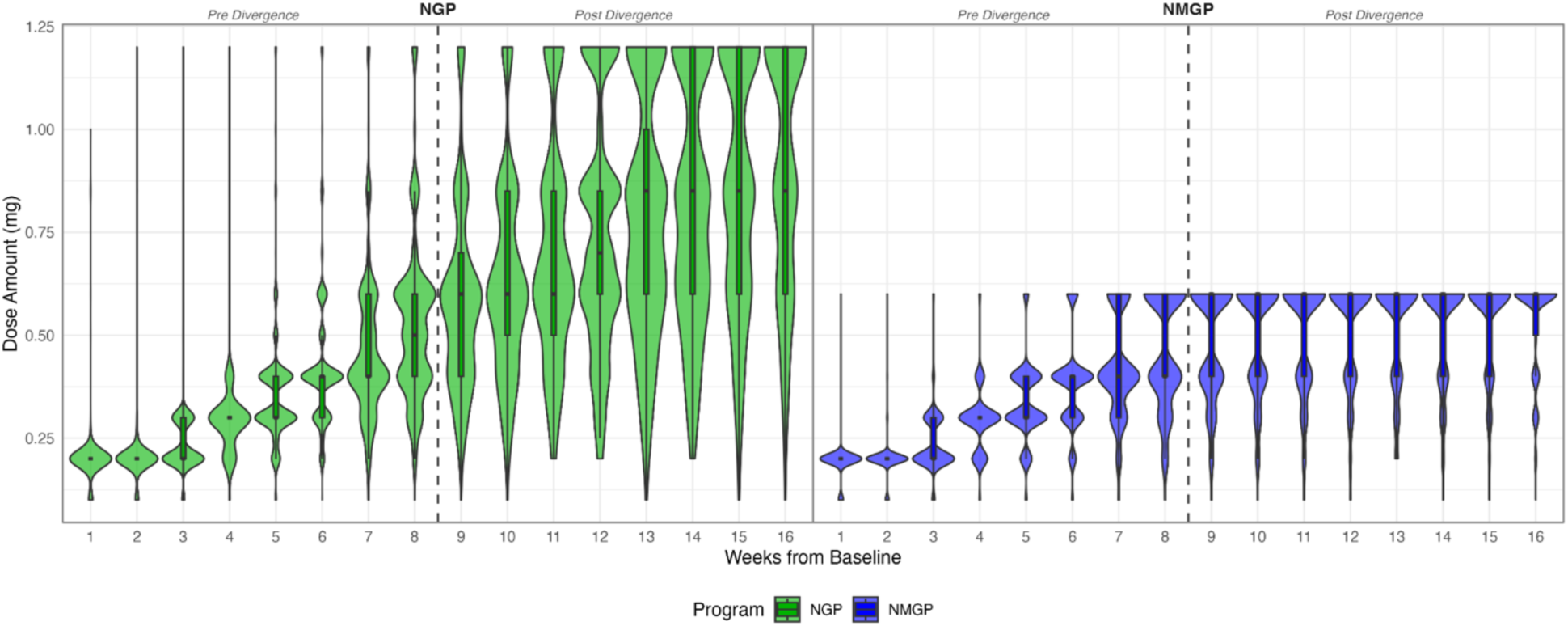
Violin plot displaying the full distribution of dose amounts each week for participants enrolled in NGP and NMGP matched cohort (n=1219 per group).

## Data Extraction and matching procedure

The data cleaning process excluded participants with insufficient weight measurements (i.e., fewer than 2 weight measurements over 16 weeks) or extreme weight change (i.e., outliers > 3 Standard Deviations), leaving 29,788 across all three Noom programs. Triplet matching via generalized propensity scores using the vecmatch R package (21) was employed, using gender, age, baseline BMI, ADI, US region, and rural versus urban RUCA categorization. For ADI, this analysis utilized the “sociome” package in R (22) in which only 1:1 matches based on ZCTA were included in the final matched cohort. Matching resulted in 3,657 participants for the matched cohort analysis. Thus, 1,219 NW participants were matched with 1,219 NMGP participants and 1,219 NGP participants. Matching quality was evaluated via standardized mean differences.

## Analytic Plan

For both the full and matched samples, weight change analyses employed a multiple-group latent growth curve model (LGCM) with a quadratic (curvature) parameter to compare longitudinal weight loss trajectories among three user groups: NW, NMGP, and NGP. The LGCM was fit using the “lavaan” package in R (23) utilizing a robust maximum likelihood estimator to ensure valid standard errors and Chi-Square statistics under potential non-normality. Full Information Maximum Likelihood (FIML) estimation was implemented to effectively handle missing data due to participant attrition, reducing bias in parameter estimates. The use of robust maximum likelihood estimation with FIML allowed inclusion of all available data under a missing-at-random assumption and provided parameter estimates that are robust to non-normal outcome distributions. For longitudinal data, FIML is reasonably able to recover parameter estimates even when attrition is high at later time points, assuming attrition can be explained by the dynamics of previous time points (24). We also examined whether missingness varied by treatment group over time.

## Results

### Baseline characteristics

The baseline date was defined as the date of the first recorded weight, and baseline characteristics for the full sample and matched samples are included in Table 1. Baseline demographics for the full and matched samples were similar, including a majority of female participants with an average age of approximately 45 years old, predominantly residing in urban areas throughout the United States, largely living in areas characterized as low or moderate in terms of deprivation. The percentage of women in the matched sample was higher than the full sample, and the percentage of participants living in areas of high deprivation was lower in the matched than the full sample.

**Table 1.** Baseline characteristics of full sample and matched cohort of compounded semaglutide program users, July 2025 - August 2025.

|  | Full Sample |  |  | Matched Cohort |  |  |
| --- | --- | --- | --- | --- | --- | --- |
| **Baseline characteristics** | **NW** | **NGP** | **NMGP** | **NW** | **NGP** | **NMGP** |
| **N, n (%)** | 20,918<br>(66.5) | 7,682<br>(24.4) | 2,877<br>(9.1) | 1,219<br>(33.3) | 1,219<br>(33.3) | 1,219<br>(33.3) |
| **Female, n (%)** | 18,128<br>(86.7) | 6,537<br>(85.1) | 2,660<br>(92.5) | 1,137<br>(93.3) | 1,141<br>(93.6) | 1,132<br>(92.9) |
| **Age, mean (SD)** | 45.28<br>(14.79) | 43.83<br>(13.25) | 44.68<br>(12.14) | 43.76<br>(14.31) | 43.18<br>(12.41) | 43.75<br>(11.91) |
| **BMI, mean (SD)** | 29.88<br>(5.29) | 32.19<br>(5.09) | 29.88<br>(4.39) | 30.37<br>(4.50) | 30.73<br>(4.33) | 30.47<br>(4.45) |
| **Northeast, n (%)** | 4,970<br>(24.1) | 1,214<br>(16.0) | 531<br>(18.7) | 179<br>(12.7) | 172<br>(12.2) | 169<br>(12.0) |
| **Midwest, n (%)** | 4,589<br>(22.2) | 1,488<br>(19.6) | 524<br>(18.4) | 191<br>(13.5) | 191<br>(13.5) | 196<br>(13.9) |
| **South, n (%)** | 5,977<br>(29.0) | 2,383<br>(31.4) | 864<br>(30.4) | 463<br>(32.8) | 479<br>(33.9) | 473<br>(33.5) |
| **West, n (%)** | 5,098<br>(24.7) | 2,493<br>(32.9) | 923<br>(32.5) | 579<br>(41.0) | 570<br>(40.4) | 574<br>(40.7) |
| **Urban, n (%)** | 17,684<br>(85.2) | 6,487<br>(85.0) | 2,489<br>(87.1) | 1,069<br>(87.7) | 1,080<br>(88.6) | 1,063<br>(87.2) |
| **Low ADI ($\leq 33$), n (%)** | 7950<br>(39.3) | 3177<br>(42.7) | 1261<br>(45.3) | 592<br>(48.6) | 608<br>(49.9) | 636<br>(52.2) |
| **Moderate ADI ($>33$ & $\leq 66$), n (%)** | 9123<br>(45.1) | 3234<br>(43.5) | 1189<br>(42.7) | 500<br>(41.0) | 484<br>(39.7) | 481<br>(39.5) |
| **High ADI ($>66$), n (%)** | 3138<br>(15.5) | 1026<br>(13.8) | 333<br>(12.0) | 127<br>(10.4) | 127<br>(10.4) | 102<br>(8.4) |

*Note.* Values are presented as n (%) for categorical variables and mean (SD) for continuous variables. NW = Noom weight; NGP = Noom GLP-1 Rx Program; NMGP = Noom Microdose GLP-1 Rx Program; ADI = Area Deprivation Index (High = High Deprivation, Low = Low Deprivation).

### Dosage distributions

Figure 1 shows the distribution of dose amounts each week for participants in the NMGP and NGP programs in the matched cohort. Although the recommended titration schedule did not differ until after week 8, it appears in both samples the NMGP and NGP average dosages started to diverge after week 5. The maximum average dosage for NGP users was approximately 0.9mg/week, whereas the maximum average dosage for NMGP users was approximately 0.5mg/week. Neither group, on average, reached the program’s defined maximum dose; however, the plot confirms NGP users, on average, took larger doses than NMGP users, and that the differences in average weekly doses diverged more over time.

### Full sample weight change (Aim 1)

Table 2, Table S1, and Figure 2 present weight change over 16 weeks in the full sample (N=29,788), prior to matching. NGP participants lost, on average, 17.3lbs, or 8.6% of their body weight by week 16, which represents significantly greater weight loss compared to NMGP participants who lost 15.0lbs, or 8.3% of their weight (Difference = 0.3% [0.1%, 0.5%], *p* = 0.001) and NW participants who lost 4.9lbs, or 2.6% of their weight (Difference = 6.0% [5.9%, 6.1%], *p* < .001). Although the NMGP and NGP trajectories appear similar (Figure 2), NMGP participants lost a significantly greater percentage of weight compared to NGP participants from weeks 1 through 7, despite identical dosing protocols. NGP participants lost a significantly greater percentage of weight compared to NMGP participants in weeks 15 and 16, despite increasing dosage at week 8. NMGP and NGP participants lost weight at a significantly greater rate each week compared to NW participants throughout the course of the study.

**Table 2.**
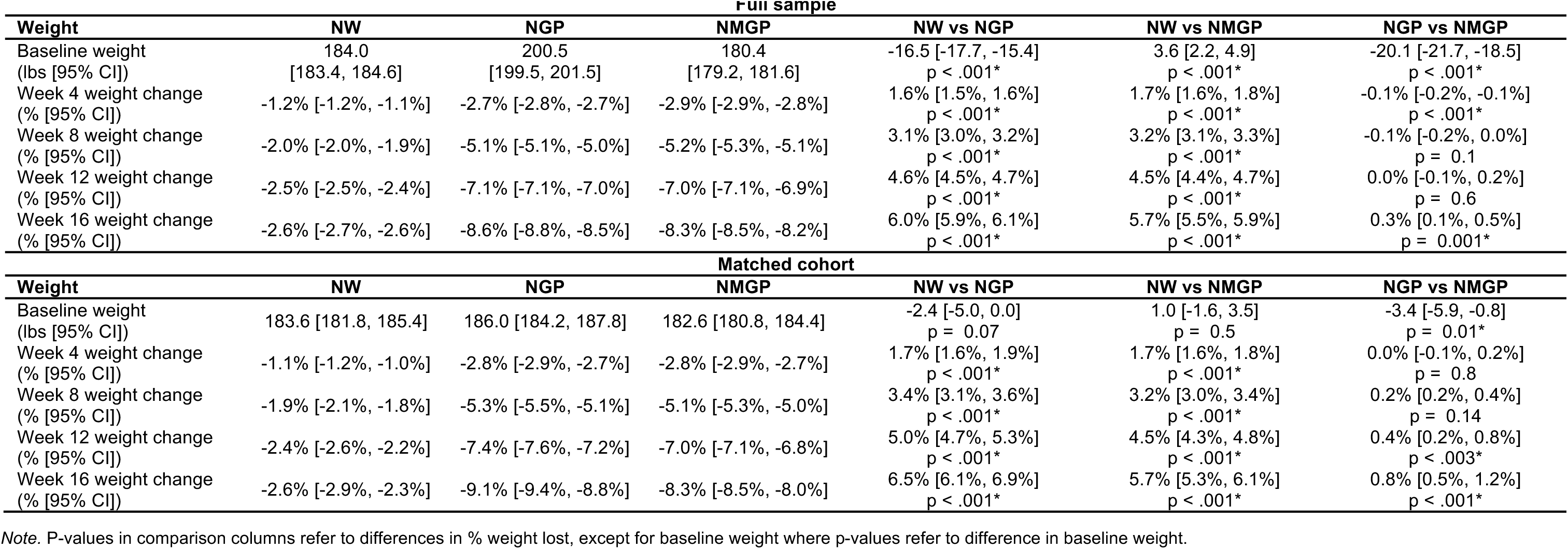
Comparison of weight loss (percentage) for full sample (N=29,788) and matched cohort (N=3657, n=1219 per group) of compounded semaglutide program users, July 2025 - August 2025.

| Full sample |  |  |  |  |  |  |
| --- | --- | --- | --- | --- | --- | --- |
| Weight | NW | NGP | NMGP | NW vs NGP | NW vs NMGP | NGP vs NMGP |
| Baseline weight (lbs [95% CI]) | 184.0<br>[183.4, 184.6] | 200.5<br>[199.5, 201.5] | 180.4<br>[179.2, 181.6] | -16.5 [-17.7, -15.4]<br>p < .001* | 3.6 [2.2, 4.9]<br>p < .001* | -20.1 [-21.7, -18.5]<br>p < .001* |
| Week 4 weight change (% [95% CI]) | -1.2% [-1.2%, -1.1%] | -2.7% [-2.8%, -2.7%] | -2.9% [-2.9%, -2.8%] | 1.6% [1.5%, 1.6%]<br>p < .001* | 1.7% [1.6%, 1.8%]<br>p < .001* | -0.1% [-0.2%, -0.1%]<br>p < .001* |
| Week 8 weight change (% [95% CI]) | -2.0% [-2.0%, -1.9%] | -5.1% [-5.1%, -5.0%] | -5.2% [-5.3%, -5.1%] | 3.1% [3.0%, 3.2%]<br>p < .001* | 3.2% [3.1%, 3.3%]<br>p < .001* | -0.1% [-0.2%, 0.0%]<br>p = 0.1 |
| Week 12 weight change (% [95% CI]) | -2.5% [-2.5%, -2.4%] | -7.1% [-7.1%, -7.0%] | -7.0% [-7.1%, -6.9%] | 4.6% [4.5%, 4.7%]<br>p < .001* | 4.5% [4.4%, 4.7%]<br>p < .001* | 0.0% [-0.1%, 0.2%]<br>p = 0.6 |
| Week 16 weight change (% [95% CI]) | -2.6% [-2.7%, -2.6%] | -8.6% [-8.8%, -8.5%] | -8.3% [-8.5%, -8.2%] | 6.0% [5.9%, 6.1%]<br>p < .001* | 5.7% [5.5%, 5.9%]<br>p < .001* | 0.3% [0.1%, 0.5%]<br>p = 0.001* |
| Matched cohort |  |  |  |  |  |  |
| Weight | NW | NGP | NMGP | NW vs NGP | NW vs NMGP | NGP vs NMGP |
| Baseline weight (lbs [95% CI]) | 183.6 [181.8, 185.4] | 186.0 [184.2, 187.8] | 182.6 [180.8, 184.4] | -2.4 [-5.0, 0.0]<br>p = 0.07 | 1.0 [-1.6, 3.5]<br>p = 0.5 | -3.4 [-5.9, -0.8]<br>p = 0.01* |
| Week 4 weight change (% [95% CI]) | -1.1% [-1.2%, -1.0%] | -2.8% [-2.9%, -2.7%] | -2.8% [-2.9%, -2.7%] | 1.7% [1.6%, 1.9%]<br>p < .001* | 1.7% [1.6%, 1.8%]<br>p < .001* | 0.0% [-0.1%, 0.2%]<br>p = 0.8 |
| Week 8 weight change (% [95% CI]) | -1.9% [-2.1%, -1.8%] | -5.3% [-5.5%, -5.1%] | -5.1% [-5.3%, -5.0%] | 3.4% [3.1%, 3.6%]<br>p < .001* | 3.2% [3.0%, 3.4%]<br>p < .001* | 0.2% [0.2%, 0.4%]<br>p = 0.14 |
| Week 12 weight change (% [95% CI]) | -2.4% [-2.6%, -2.2%] | -7.4% [-7.6%, -7.2%] | -7.0% [-7.1%, -6.8%] | 5.0% [4.7%, 5.3%]<br>p < .001* | 4.5% [4.3%, 4.8%]<br>p < .001* | 0.4% [0.2%, 0.8%]<br>p < .003* |
| Week 16 weight change (% [95% CI]) | -2.6% [-2.9%, -2.3%] | -9.1% [-9.4%, -8.8%] | -8.3% [-8.5%, -8.0%] | 6.5% [6.1%, 6.9%]<br>p < .001* | 5.7% [5.3%, 6.1%]<br>p < .001* | 0.8% [0.5%, 1.2%]<br>p < .001* |

Note. P-values in comparison columns refer to differences in % weight lost, except for baseline weight where p-values refer to difference in baseline weight.

**Figure 2.**
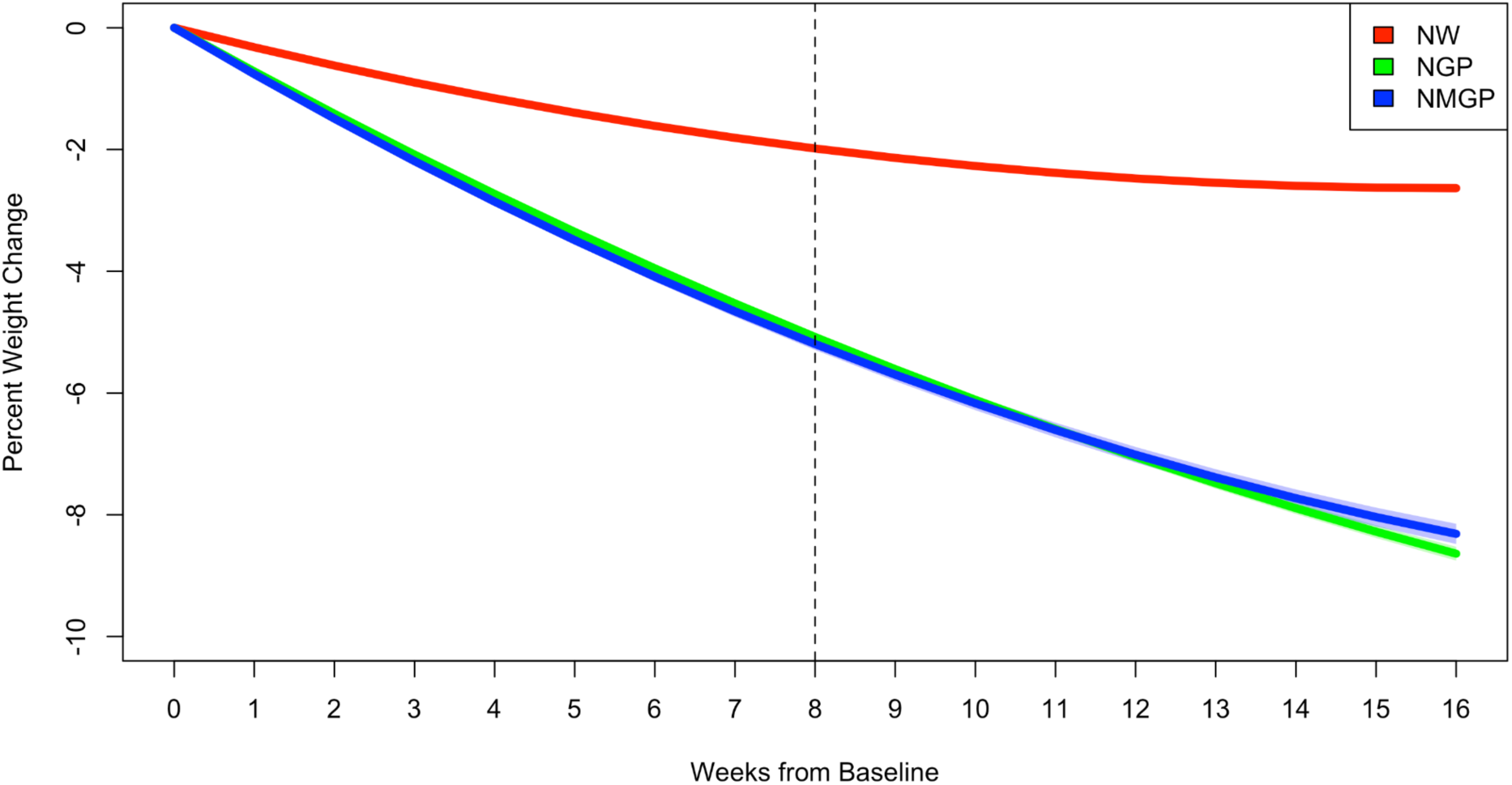
Estimated latent growth curve trajectories of percent weight loss for full sample with identical titration protocols for the first 8 weeks of program enrollment before titration cap of 0.6mg/weekly for NMGP and continuing titration up to 1.2mg/weekly for NGP in weeks 9-16 (N=29,788).

A chi-square test comparing the proportion of missingness between groups over time was non-significant (χ²(7) = 8.23, *p* = .313), indicating no meaningful differences in missing data patterns by treatment group.

### Matched sample weight change (Aim 2)

Post-matching data shows acceptable matching on all matching variables (see Figure 3). Table 2, Table S1, and Figure 4 present weight change over 16 weeks in the matched sample (N= 3,657; n=1,219 per program). Matched NGP participants lost, on average, 16.9lbs, or 9.1% of their body weight by week 16, which represents significantly greater weight loss compared to NMGP participants who lost 15.1lbs, or 8.3% of their weight (Difference = 0.8% [0.5%, 1.2%], *p* < 0.001) and NW participants who lost 4.8lbs, or 2.6% of their weight (Difference = 6.5% [6.1%, 6.9%], *p* < 0.001). Although the NMGP and NGP early trajectories appear similar (Figure 4), NGP participants lost a significantly greater percentage of weight compared to NMGP participants in weeks 9 through 16, which largely corresponds with the increased dosage at week 8 for NGP users. NMGP and NGP participants lost weight at a significantly greater rate each week compared to NW participants throughout the course of the study.

**Figure 3.**
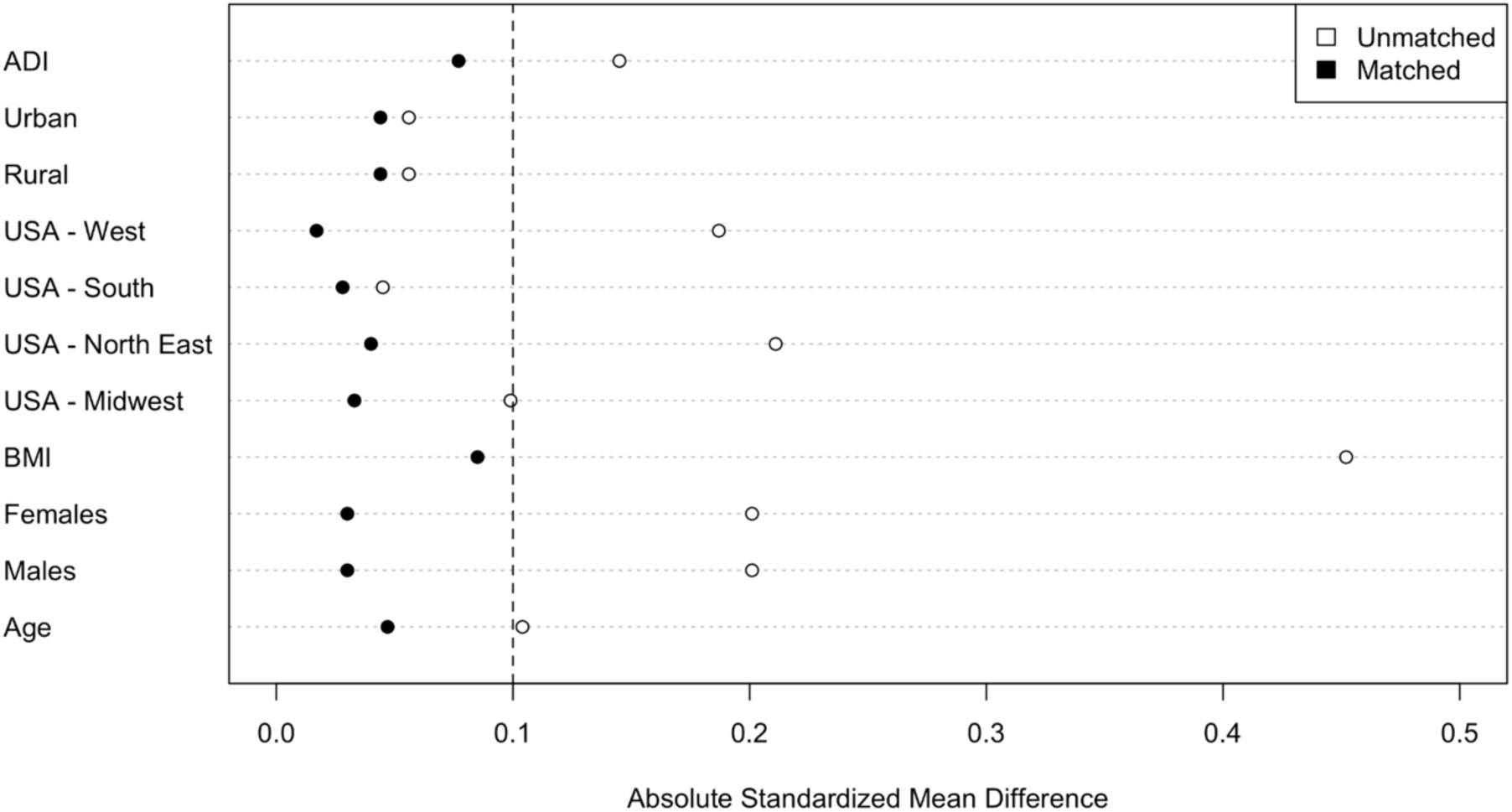
Matching quality, pre- and post-matching results. Post-matching data shows acceptable matching on all matching variables (N=3657, n=1219 per group).

**Figure 4.**
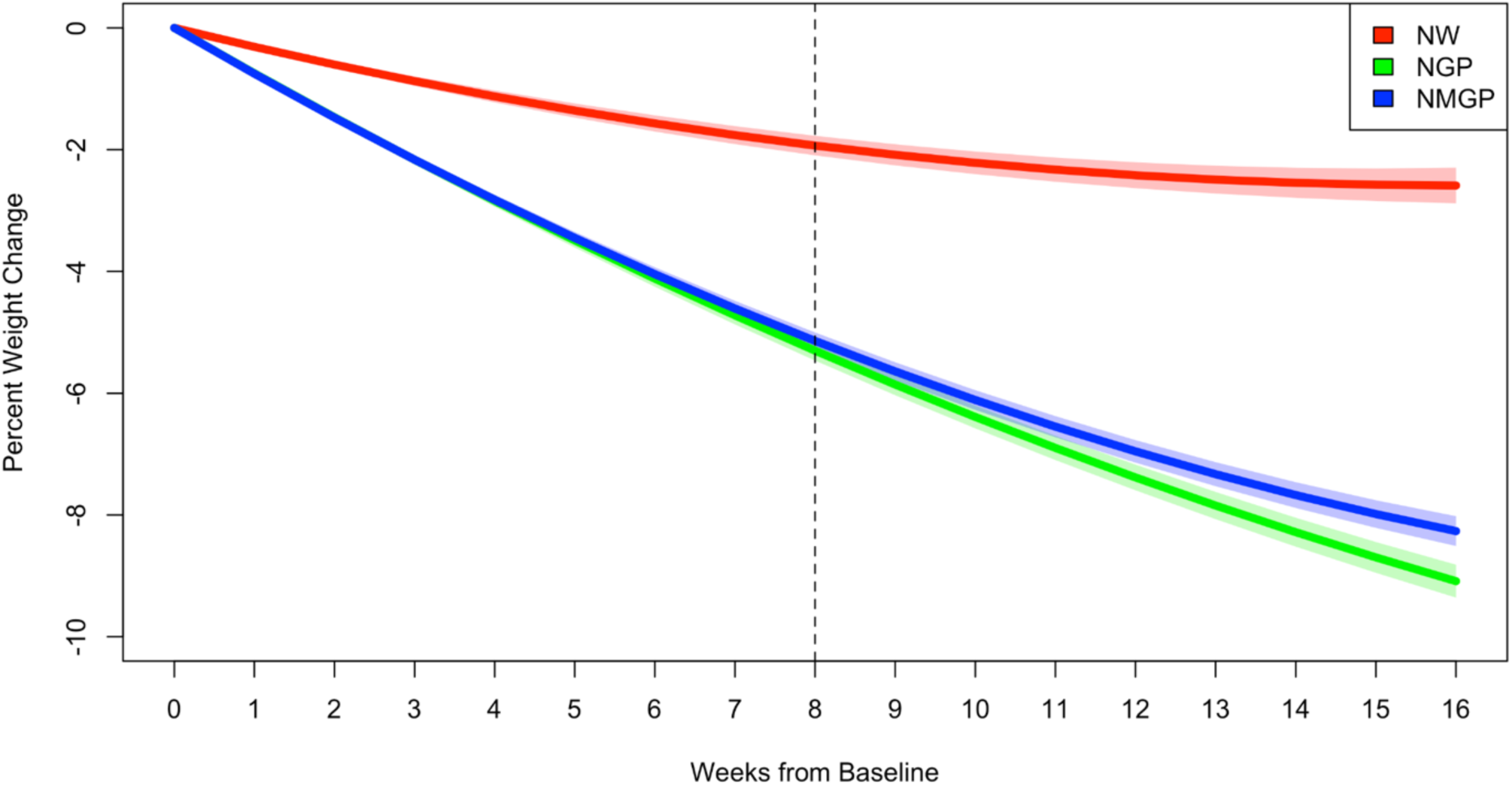
Estimated latent growth curve trajectories of percent weight loss for matched cohort with identical titration protocols for the first 8 weeks of program enrollment before titration cap of 0.6mg/weekly for NMGP and continuing titration up to 1.2mg/weekly for NGP in weeks 9-16 (N=3657, n=1219 per group).

## Discussion

In this retrospective, real-world study, results revealed NMGP users who were prescribed lower doses of compounded semaglutide (up to 0.6mg/weekly) lost almost as much weight in the 16 weeks after initiating treatment as NGP users who could titrate up to Noom’s standard dose of compounded semaglutide (up to 1.2mg/weekly). In fact, in the full sample, the difference in percentage of weight lost at 16 weeks between microdose (NMGP) and Noom’s standard dose (NGP) users was 0.3%. In the matched cohort, NMGP users achieved roughly 91% of the weight loss observed with NGP, equating to approximately 1 pound per week in both groups. Clinically significant weight loss (>5%) was observed among NMGP and NGP users, with mean weight loss of over 8% for NMGP and NGP users at week 16 in the full and matched samples. Results of the present study support a philosophy of titrating to the minimum effective dose of compounded semaglutide to achieve desired outcomes.

Weight loss among NMGP users in the current study appears to be similar to weight loss observed in a study of lower dose, FDA approved semaglutide conducted over a similar timeframe (11). In the present study, NMGP users lost, on average, 12.7lbs in 12 weeks and 15lbs at 16 weeks taking a compounded semaglutide. In comparison, lower dose FDA approved semaglutide users in Nauck and colleagues (11) lost 4.9kg (10.7lbs) over 12 weeks. This comparison has important implications for the approximate 20% of GLP-1 users who take compounded formulations (3). Unlike Nauck and colleagues (11), in the present study, participants were engaged in a behavioral weight management program alongside medication treatment (i.e., Noom companion experience with mobile application, Success Kit, and medication management). Pairing a GLP-1 with a behavioral weight management program has been previously reported to optimize weight loss (9,15).

Despite taking, on average, approximately half the dose of compounded semaglutide by week 16 as NGP users, NMGP users demonstrated comparable weight loss over the observed treatment period. In the full sample, the NGP group did not produce significantly higher weight loss percentage until week 15. These results suggest either the increase in dose took time to accelerate weight loss in the NGP group or perhaps engagement in behavioral modification differed between groups. Lower doses of compounded semaglutide may be an option for patients with cost barriers to higher dose FDA approved or compounded formulations (5,25). In addition, lower doses of FDA approved semaglutide have been associated with fewer adverse events and improved tolerability (11). In the present study, NMGP users had up to 38% lower odds of reporting any side effects by week 16 compared to NGP users (26). The combination of similar early term weight loss and reduced side effects (26) suggest a lower dose (up to 0.6mg/week) of compounded semaglutide shows promise as a weight management solution when paired with a digital behavioral companion.

Matched cohort analyses were conducted to add methodological rigor, mimicking quasi-experimental comparisons, to substantiate results in the full sample. Results of matched analyses were similar to those of the full sample, thus results held when methodological rigor was enhanced. There were slight differences in results of analyses for the full versus matched samples, with a slightly greater percentage weight loss observed among NGP users in the matched cohort compared to the full sample. This may be due to the nature of matching, wherein fewer male than female NGP users were able to be matched, reducing baseline weight by almost 15lb. Despite this difference, absolute weight loss was similar (17.3lb and 16.9lb), thus weight loss represented a somewhat larger proportion of baseline body weight in the matched NGP cohort (9.1%) than the full sample of NGP users (8.6%). Last, this study’s endpoints (week 16) were measured in November and December. Prior research indicates weight gain is typically observed during these months (27).

## Limitations

There are several important limitations to the present study that future research should seek to address. First, the full sample was largely female, and the matched sample consisted of an even larger proportion of urban women, limiting the generalizability of results. However, the demographic composition of the sample is broadly consistent with the characteristics of users taking FDA approved GLP-1 medications reported in real-world studies (28). Because this was a real-world study rather than an RCT, compounded semaglutide dosing was not experimentally controlled. Although there was a clinical protocol, dosing was personalized based on participants’ weight loss response and tolerability, resulting in some variation in dose and timing of dose escalation. For example, some NGP participants exceeded the 0.6mg interim ceiling before the protocol-specified increase at week 9. Dosing patterns increasingly diverged between NGP and NMGP participants over time (Figure 1). Accordingly, observed outcomes reflect treatment as implemented in clinical practice rather than the effects of strict, standardized dosing regimens.

As retrospective real-world study, it utilized a convenience sample, meaning participants were not randomized but rather selected based on their enrollment in NW programs. To partially address this, analyses included descriptives of the sample and matching was employed to statistically control for measured baseline characteristics that could influence weight loss, thereby strengthening the internal validity of the study. While ADI has been shown to be a useful proxy for the socioeconomic disparities, even matching at the ZCTA level does not explicitly incorporate race, ethnicity, or socioeconomic status (29). Area-level deprivation indices may be sensitive to both the geographic level at which they are calculated and their underlying construction (29). While ZCTA-level measures provide a useful proxy when individual-level socioeconomic data are unavailable, they represent relatively large geographic areas and may obscure within-ZCTA heterogeneity, resulting in residual confounding or imprecise representation of participants’ individual socioeconomic circumstances (30). To minimize potential misclassification, only ZIP codes with a one-to-one ZCTA match were included in the matched cohort.

While Noom users with Type 2 diabetes were excluded from the present study, other medical comorbidities (beyond inclusion/exclusion criteria) were not measured and thus could not be used for matching purposes. Unmeasured confounding from these unmatched variables could skew treatment effect estimates. Additionally, body weight and medication dosage were obtained by self-report, which may introduce bias via measurement error. Prescription fills, medication logs, or actual doses administered were not verified.

The study also experienced participant drop-out, which raises concerns about attrition bias, where the loss of participants is not random but related to treatment assignment, cost, or outcome, and remaining participants may disproportionately represent those who responded well to treatment, leading to survivor bias and an overestimation of effects. FIML was used in analyses to account for attrition. In addition, this population consists of early adopters to a lower dose program who self-selected enrollment at program launch. Early adopters may be systematically different from later adopters in characteristics such as motivation or patient activation. Finally, it is not known whether weight loss results of NMGP can be generalized to other compounding semaglutide manufacturers given potential variability in exact formulations, or if results translate to FDA approved GLP-1 medications.

This study was conducted by a team of researchers employed by Noom, Inc. We support and recommend external validation of our findings to strengthen confidence in these results and further research to evaluate the safety, tolerability, and efficacy of compounded semaglutide.

## Conclusions

This large, retrospective study of Noom users enrolled in three programs (NW, NMGP, and NGP) revealed lower doses of compounded semaglutide (up to 0.6mg/weekly) produced 91% of the weight loss achieved by higher compounded GLP-1 doses (up to 1.2mg/weekly) in the first sixteen weeks of treatment. Analyses were conducted on the full sample as well as a matched cohort. Results of both revealed NMGP and NGP users, on average, lost over 8% of their baseline weight at 16 weeks, a clinically significant reduction in weight. When paired with a behavioral weight management program companion, lower doses of compounded semaglutide show significant promise to deliver clinically meaningful weight loss.

## Supplementary Material

***Table S1.*** Comparison of weight loss (lbs) for full sample and matched cohort of compounded GLP-1 RA program users, July 2025 - August 2025.

## Clinical trial registration

N/A

## Disclosure

Authors E.C.O., R.G. M., M.C.A., and A.F. are employees at Noom, Inc. and have received salary and stock or stock options for their employment. Authors J.L.M. and A.R.M. are contractors at Noom, Inc. and have received payment for their services.

## Data Availability

All data produced in the present study are available upon reasonable request to the authors.

## Acknowledgments

The authors thank Whitney Evans for her thoughtful feedback on an earlier draft of this manuscript.

## Author Contribution

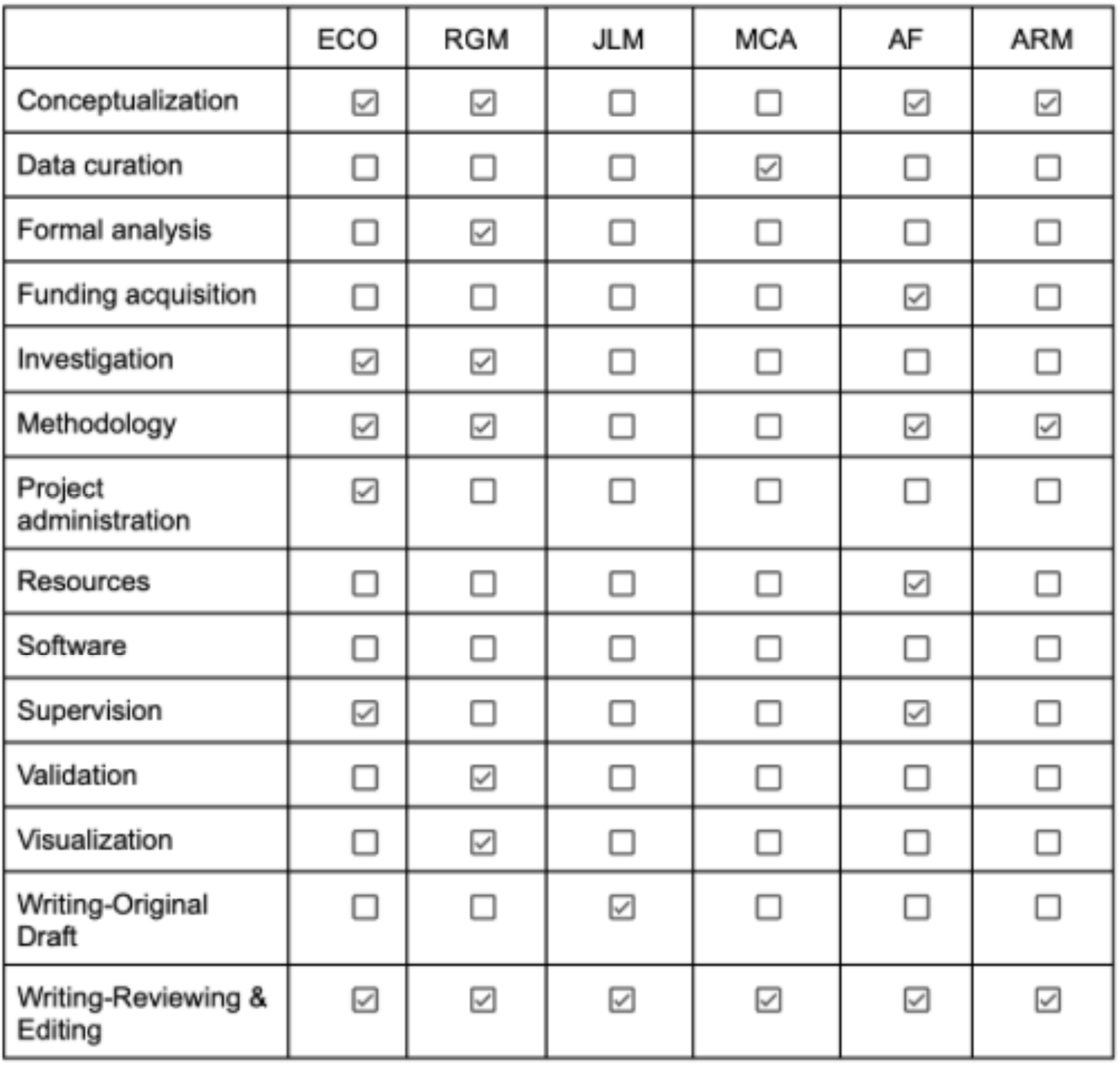

