## Supplementary materials for "Weight loss with a lower dose compounded semaglutide and behavioral weight management program: A real-world matched retrospective cohort study"

***Table S1.*** Comparison of weight loss (lbs) for full sample (N=29,788) and matched cohort (N=3657, n=1219 per group) of compounded semaglutide program users, July 2025 - August 2025.

| | | | Full sample | | | |
|---|---|---|---|---|---|---|
| **Weight** | **NW** | **NGP** | **NMGP** | **NW vs NGP** | **NW vs NMGP** | **NGP vs NMGP** |
| Baseline weight (lbs [95% CI]) | 184.0 [183.4, 184.6] | 200.5 [199.5, 201.5] | 180.4 [179.2, 181.6] | -16.5 [-17.7, -15.4], p < .001* | 3.6 [2.2, 4.9], p < .001* | -20.1 [-21.7, -18.5], p < .001* |
| Week 4 weight change (lbs [95% CI]) | -2.1 [-2.2, -2.1] | -5.5 [-5.6, -5.4] | -5.2 [-5.3, -5.1] | 3.3 [3.3, 3.4], p < .001* | 3.0 [2.9, 3.1], p < .001* | 0.3 [0.2, 0.5], p < .001* |
| Week 8 weight change (lbs [95% CI]) | -3.7 [-3.8, -3.6] | -10.2 [-10.3, -10.1] | -9.4 [-9.6, -9.2] | 6.5 [6.4, 6.7], p < .001* | 5.7 [5.5, 5.9], p < .001* | 0.8 [0.6, 1.0], p < .001* |
| Week 12 weight change (lbs [95% CI]) | -4.6 [-4.7, -4.5]; | -14.1 [-14.3, -14.0] | -12.7 [-12.9, -12.4] | 9.6 [9.4, 9.8], p < .001* | 8.1 [7.9, 8.4], p < .001* | 1.5 [1.2, 1.8], p < .001* |
| Week 16 weight change (lbs [95% CI]) | -4.9 [-5.0, -4.7] | -17.3 [-17.6, -17.1] | -15.0 [-15.3, -14.7] | 12.5 [12.2, 12.7], p < .001* | 10.2 [9.8, 10.5], p < .001* | 2.3 [2.0, 2.7], p = 0.001* |
| | | | **Matched cohort** | | | |
| **Weight** | **NW** | **NGP** | **NMGP** | **NW vs NGP** | **NW vs NMGP** | **NGP vs NMGP** |
| Baseline weight (lbs [95% CI]) | 183.6 [181.8, 185.4] | 186.0 [184.2, 187.8] | 182.6 [180.8, 184.4] | -2.4 [-5.0, 0.0], p = 0.07 | 1.0 [-1.6, 3.5], p = 0.5 | -3.4 [-5.9, -0.8], p = 0.01* |
| Week 4 weight change (lbs [95% CI]) | -2.1 [-2.3, -1.19] | -5.3 [-5.5, -5.1] | -5.2 [-5.3, -5.0] | 3.2 [3.0, 3.5], p < .001* | 3.1 [2.9, 3.3], p < .001* | 0.1 [-0.1, 0.4], p = 0.3 |
| Week 8 weight change (lbs [95% CI]) | -3.6 [-3.9, -3.3] | -9.9 [-10.2, -9.6] | -9.4 [-9.7, -9.1] | 6.3 [5.9, 6.7], p < .001* | 5.8 [5.5, 6.2], p < .001* | 0.5 [0.1, 0.9], p = 0.02* |
| Week 12 weight change (lbs [95% CI]) | -4.4 [-4.8, -4.1] | -13.7 [-14.1, -13.3] | -12.7 [-13.0, -12.4] | 9.3 [8.7, 9.9], p < .001* | 8.3 [7.7, 8.8], p < .001* | 1.0 [0.5, 1.6], p < .001* |
| Week 16 weight change (lbs [95% CI]) | -4.8 [-5.3, -4.2] | -16.9 [-17.4, -16.4] | -15.1 [-15.6, -14.6] | 12.2 [11.4, 12.9], p < .001* | 10.3 [9.6, 11.0], p < .001* | 1.8 [1.1, 2.5], p < .001* |

*Note.* P-values in comparison columns refer to differences in weight lost (lbs).
